# When Is Maltreatment ‘Acceptable’? Cross-Cultural Patterns in Child Maltreatment Exposure and its Perceived Social Acceptability Across 13 Countries and Regions

**DOI:** 10.64898/2026.10.07.26364967

**Authors:** Dan Jenkins, Rachel Langevin, Stefanie Rita Balle, O Jiaqing, Nino Makhashvili, Shir Porat-Butman, Einat Levi-Gigi, Görkem Ayas, Deniz Ceylan, Fares-Otero Natalia, Mónica Fernández Boente, Halligan Sarah, Billy Jansson, Saman Kamari Songhorabadi, Ghasem Khayati, Farhad Khormaei, Chantal Martin-Sölch, Monique Pfaltz, Mayron Piccolo, Vedat Şar, Ulrich Schnyder, Soraya Seedat, Tanya Tandon, Tuğba Türk Kurtça, Eleonora Bartoli, Georgina Spies, Naved Iqbal, Misari Oe, CCTS Child Trauma Network

**Affiliations:** Department of Psychiatry, Stellenbosch University, South Africa; Department of Psychology, University of Fribourg, Switzerland; Department of Educational and Counselling Psychology, McGill University, Canada; Department of Psychology, LMU Munich, Germany; Department of Psychology, University of Macau, Macau; Ilia State University, Tbilisi, Georgia; Faculty of Education, Bar-Ilan University, Israel; Gonda Multidisciplinary Brain Research Center, Bar-Ilan University, Israel; Koç University Graduate School of Health Sciences, Istanbul, Türkiye; Department of Psychiatry and Psychology, Mayo Clinic, Rochester, MN, USA; Department of Psychiatry, School of Medicine, Koç University, Istanbul, Türkiye; Koç University Research Center for Translational Medicine (KUTTAM), Affective Laboratory, Istanbul, Türkiye; Department of Psychiatry and Psychotherapy, Technical University of Munich (TUM), Munich, Germany; Department of Psychology, University of Bath, Bath, UK; Department of Education, Psychology and Social Work, Mid Sweden University, Sweden; Faculty of Educational Sciences and Psychology, Shiraz University, Shiraz, Iran; Department of Psychology and Cognitive Science, University of Trento, Rovereto, Italy; Institute for Cognitive Neuroscience, Faculty of Social Sciences, Higher School of Economics, Moscow, Russian Federation; Department of Psychology, Harvard University, USA; Department of Psychiatry, Harvard Medical School, USA; University of Zurich, Switzerland; Department of Psychological Counseling and Guidance, Faculty of Education, Trakya University, Edirne, Türkiye; Institute of Psychology, Goethe University, Frankfurt am Main, Germany; Department of Psychology, University of Cape Town, South Africa; Department of Psychology, Jamia Millia Islamia, Delhi, India; Department of Neuropsychiatry, Kurume University School of Medicine, Japan

**Keywords:** childhood trauma, cross-cultural, social norms, mental health, resilience, psychological distress

## Abstract

Child maltreatment (CM) is a global phenomenon, yet its prevalence, acceptability, and psychosocial consequences vary across cultures. This study examined self-reported CM, its perceived acceptability, and associations with psychosocial outcomes among 1,617 adults (*M*age = 25.21, *SD* = 6.74) across 13 countries and regions. Participants completed an online survey assessing physical neglect, physical abuse, emotional maltreatment, sexual abuse, and witnessing domestic violence, alongside perceived acceptability, post-traumatic growth, resilience, perceived social support, post-traumatic symptoms, and psychological distress. Country/region differences were tested via mixed-effects; whether CM–outcome associations varied by country/region was tested via fixed-effects interaction models. Significant cross-country/region variability emerged for most CM subtypes, especially for emotional maltreatment. Witnessing domestic violence rates did not differ significantly, though its acceptability still varied significantly, as did all other types. Greater CM exposure was linked to poorer mental health for several subtypes and to greater acceptability of maltreatment, raising concerns about intergenerational normalisation. CM–outcome associations were largely consistent across countries/regions, with only isolated, exploratory exceptions. Findings show that while CM’s adverse psychosocial impact shows little cross-national variation, its prevalence and normative framing are culturally shaped, underscoring the need for culturally sensitive assessment and intervention.

**Impact statement:** Child maltreatment, including physical and emotional abuse, neglect, sexual abuse, and witnessing domestic violence, affects children in every part of the world, yet most research on its effects comes from a small number of high-income, Western countries. This study surveyed 1,617 adults across 13 countries and regions spanning five continents, placing it among the most culturally diverse investigations to date of how child maltreatment is experienced, perceived, and linked to mental health.

The findings carry two practical messages. First: how common maltreatment is and how socially acceptable people consider it to be differs substantially across countries and regions. Policymakers, educators, and child-protection professionals who design and implement prevention and awareness efforts cannot rely on a single global template; they need to adapt to local norms around parenting and discipline. Second: the psychological harm caused by maltreatment, i.e., poorer mental health and greater psychological distress, was remarkably consistent across all 13 settings. This suggests that regardless of a culture’s attitudes toward specific parenting practices, the damage maltreatment does to children’s wellbeing is a shared human experience, reinforcing the case for maltreatment prevention as a universal child health priority rather than a culturally relative one.

The study also found that people who experienced more maltreatment as children were more likely to view it as acceptable, raising concerns that abusive practices may be passed down between generations through shifting social norms, not only through the direct repetition of behaviour. This suggests an avenue for intervention: efforts that address attitudes and beliefs about discipline and family life, not just behaviour, may help break cycles of harm. Together, these findings can inform the design of culturally responsive screening tools, clinical training, and public health campaigns aimed at protecting children worldwide.

## 1. Introduction

Child maltreatment (CM), encompassing physical and sexual abuse, emotional maltreatment, neglect, and witnessing domestic violence before age 18, affects between 12.7% and 36.3% of children worldwide, depending on the type and region (Moody *et al*. 2018). CM is consistently linked to compromised physical and mental health and difficulties in social relationships (Hughes *et al*. 2017; Pfaltz *et al*. 2022), although some survivors display resilient functioning and post-traumatic growth (Fares-Otero *et al*. 2025). These outcomes are not uniform across cultures: cultural norms shape how individuals process and respond to events (Mbagaya *et al*. 2013), and societal attitudes towards parenting shape both CM prevalence and how survivors make sense of their experiences (Ganz *et al*. 2020; Klika *et al*. 2019).

### 1.1 Exposure to and perceived acceptability of child maltreatment worldwide

Global CM prevalence varies considerably by type, region, and gender, with physical abuse and neglect most common in Africa and South America, and sexual and emotional abuse most frequently reported by girls in North America and Australia (Moody *et al*. 2018). Measuring and comparing CM rates across cultures is challenging because parenting practices are cultural embedded (Bornstein 2013; Lansford 2010): practices such as corporal punishment may be normative in some contexts but considered abusive in others (Cuartas *et al*. 2019; Klassen *et al*. 2022; Zolotor and Puzia 2010), and cultural norms around shame and patriarchy can limit disclosure of sexual abuse and intimate partner (Fontes and Plummer 2010; Kolhatkar and Berkowitz 2014). Our pilot study found that, although perceived acceptability of CM was generally low across countries, there was cross-country variation (Bartoli *et al*. 2024; Wadji *et al*. 2023b), highlighting the need for culturally sensitive approaches to risk assessment and event interpretation.

### 1.2 Maladaptive outcomes of child maltreatment across cultures

Meta-analytic evidence consistently links CM to a range of negative outcomes including impaired physical and cognitive development, maladaptive attachment, and elevated risk of psychological and social difficulties such as anxiety disorders, post-traumatic stress disorder (PTSD), poverty, and homelessness (Carr *et al*. 2020a, 2020b). The intensity of these outcomes appears to vary between cultures: Asians who experience CM tend to report poorer self-esteem than similarly exposed North Americans (Zhang *et al*. 2023); and the CM-PTSD association, while consistent across cultures, differs in strength by socio-economic context (Hashim *et al*. 2026). For example, physical abuse predicted mental health problems among Kenyan and Zambian, but not Dutch, participants (Mbagaya *et al*. 2013), and our pilot study similarly found that the CM-PTSD relationship varied by country and CM type, with significant associations in Cameroon and Germany but not Japan or Canada (Bartoli *et al*. 2024).

### 1.3 Positive adaptation following child maltreatment across cultures

Recovery from CM exposure has been linked to post-traumatic growth (PTG; Jankovic *et al*. 2022); resilience (Fares-Otero *et al*. 2025); and perceived social support (Evans *et al*. 2013). Findings on PTG after CM are mixed and appear to depend on CM type: childhood neglect, but not abuse, has been linked to poorer PTG outcomes (Chen *et al*. 2025), and traumatic childhood experiences can predict lower PTG directly while also showing an indirect positive association via acceptance (Quan *et al*. 2022). In contrast, our prior cross-cultural study found CM positively associated with PTG, with the highest levels observed in Cameroon, where CM was most frequent (Wadji *et al*. 2023a).

Resilience functions as a culturally shaped process rather than a fixed trait (Ungar 2013; Yoon *et al*. 2021): components such as perseverance and social support appear universal, but cultures differ in how these are realised and valued (Ungar 2013) and cultural context may moderate whether CM-related changes are viewed as strengths or weaknesses (Yu *et al*. 2022). In our pilot study, resilience scores were significantly lower in Japan than in other countries (Wadji *et al*. 2023a), and a meta-analysis similarly found stronger associations between emotional abuse and emotion regulation, but weaker associations with self-esteem, in Western versus non-Western countries (Fares-Otero *et al*. 2025).

While CM survivors often report feeling isolated and lacking support from significant others (De Heer *et al*. 2024; Pigeon *et al*. 2025), perceived social support has consistently shown a buffering effect against CM across cultures (Evans *et al*. 2013). Given culture’s central role in shaping resilience, social support, and PTG, a cross-cultural investigation of their interplay with CM and its acceptability is warranted.

### 1.4 The current study

The current study builds on a smaller-scope study conducted in Canada, Cameroon, Japan, and Germany (Bartoli *et al*. 2024; Wadji *et al*. 2023a, 2023b) which showed that rates and perceived acceptability of CM differed across countries and that culture may shape individual responses to trauma. This past study was limited by small samples and few countries and did not offer a comprehensive picture of psychosocial adaptation among CM-exposed individuals. Moreover, comparatively little is known about how perceived acceptability of CM varies across cultures and how this relates to survivors’ outcomes.

To fill these gaps, the current study explores self-reported rates of CM exposure and perceived acceptability of CM, and their associations with negative and positive mental health outcomes, among adults from 13 countries and regions representing a variety of cultures (e.g., collectivistic vs. individualistic, varied importance of religion and tradition), languages, and socioeconomic realities (i.e., high-income and low-to middle-income countries, varied perpetration of violence, and differing levels of wealth inequality). A registered study protocol (https://osf.io/ncq4r) guided the work, and ten research questions were developed before the study began, although the protocol was published after data collection. This manuscript addresses the first three questions through a cross-sectional online survey: (1) do self-reported rates of CM differ across countries and regions; (2) do countries and regions differ regarding perceived acceptability of CM; and (3) does the relationship between CM and (a) negative mental health outcomes and (b) positive adaptation vary across countries and regions?

## 2. Methods

### 2.1 Sample

We administered an online questionnaire to the general population in Canada (*n* = 180), Georgia (*n* = 101), Germany (*n* = 95), India (*n* = 136), Iran (*n* = 94), Israel (*n* = 85), Macau (*n* = 127), South Africa (*n* = 139), Sweden (*n* = 100), Switzerland (*n* = 110), Türkiye (*n* = 134), the United Kingdom (*n* = 206) and the United States (*n* = 113). Inclusion criteria were individuals aged 18–40 residing in one of these countries and regions; a range selected to isolate the developmental window where psychological shifts and social vulnerabilities intersect as key drivers of lifelong health and social disparities (Rod *et al*. 2025). Our final sample comprised 1,617 adults (*M*_age_ = 25.21, *SD* = 6.74).

### 2.2 Procedure

Advertisements were distributed on social media (Facebook, Instagram), campus message boards or departmental subject pool systems, or circulated among researchers’ personal and professional networks. Incentives differed by country (see Table S1). We administered the questionnaire online using the Stellenbosch University REDCap platform (Harris *et al*. 2019) in English, Farsi, French, Georgian, German, Hebrew, Swedish, or Turkish (see Table S2 for translation details). Participants completed an informed consent document and screening questions before accessing the study questionnaires. Helpline and counselling contact information were displayed in the consent document and above sensitive questions. Umbrella approval was granted by the Stellenbosch University Health Research Ethics Committee (ref: N23/03/026), with each partnering university providing further local ethics clearance. The project formed part of the Global Collaboration on Traumatic Stress (GCTS; Olff *et al*. 2020) child trauma network.

### 2.3 Instruments

#### 2.3.1 Sociodemographic information

Participants reported age, gender, country/region of birth and residence (and period of residence if different from birthplace), relationship status, number of children, ethnicity, years of formal education, and disability status, as well as level of contact and closeness with family members.

#### 2.3.2 Child maltreatment

CM experiences were assessed using a 24-item self-report measure of physical and sexual abuse, emotional maltreatment, physical neglect, and witnessing domestic violence. Abuse was assessed with the Early Trauma Inventory Self-Report Short Form (ETI-SR-SF; Bremner *et al*. 2007): five items for physical abuse (KR-20 = .69), five for emotional maltreatment (KR-20 = .87), and six for sexual abuse (KR-20 = .82), each summed to a subscale score (range = 0–5 for physical abuse and emotional maltreatment; 0–6 for sexual abuse). Physical neglect was assessed using five yes/no items from the ISPCAN Child Abuse Screening Tool (ICAST; Zolotor *et al*. 2009), summed across affirmative responses (range = 0–5; KR-20 = .60). Exposure to domestic violence was assessed using a three-item adaptation of the Revised Conflict Tactics Scale 2 (Straus *et al*. 1996; range = 0–3; α = .75).

#### 2.3.3 Perceived acceptability of CM

Items assessing perceived social acceptability of CM were derived from the CM items (see Wadji *et al*. 2023b for adaptation details). Participants rated how acceptable most people in their community would find each described behaviour on a five-point scale (1 = never acceptable, 5 = always acceptable), with lower scores indicating lower perceived acceptability. The adapted ICAST (α = .83), CTS2 (α = .89), and ETI-SR-SF (physical abuse α = .88, emotional maltreatment α = .94, sexual abuse α = .94) showed good internal reliability.

#### 2.3.4 Positive adaptation

Post-traumatic growth was assessed using the Post-traumatic Growth Inventory, Short-Form (PTGI-SF; Cann *et al*. 2010), a 10-item measure of adaptation following trauma exposure (α = .90) across five domains: relating to others, new possibilities, personal strength, spiritual change, and appreciation of life; rated from 1 (did not experience this change) to 6 (experienced this change to a very great degree). Responses were averaged to give a total score and five subscale scores (range 1–6). Resilience was measured using the Brief Resilience Scale (BRS; Smith *et al*. 2008), a 6-item measure of ability to recover from stressful or traumatic events (α = .83) rated on a 1–5 Likert scale (reverse-scored for negatively worded items) and averaged to a total score (range 1–5) with higher scores indicating greater resilience. Perceived social support was measured with the Multidimensional Scale of Perceived Social Support (MSPSS; Zimet *et al*. 1988), a 12-item scale assessing support from significant others, family, and friends (4 items each; total α = .92), rated on a 1–7 Likert scale and averaged for the total and each subscale (range 1–7).

#### 2.3.5 Negative mental health outcomes

Post-traumatic symptoms were assessed using the Global Psychotrauma Screen (GPS; Olff *et al*. 2021) comprising 17 yes/no symptom items and 5 yes/no protective factor-items summed to give scale scores. Participants were asked to think of the traumatic event currently affecting them most and to comment on its characteristics and timing. Beyond the 17-item total score (KR-20 = .90), the GPS includes subscales for post-traumatic stress disorder, disturbances in self-organisation, anxiety, depression, insomnia, self-harm, dissociation, and substance use, with higher subscale scores indicating more severe symptoms. Psychological distress was measured with the Kessler Psychological Distress Scale (K6; Kessler *et al*. 2003), a 6-item non-specific distress screener (α = .87), rating the frequency of negative emotions over the past 30 days relative to participants’ normal state.

### 2.4 Data analysis

Analyses were conducted in R (version 4.4.1; R Core Team 2026) using the *lme4* package (Bates 2010) for mixed-effects modelling. We addressed each research question using nested linear models comparing between-country/region variance in CM, perceived acceptability of CM, and psychosocial outcomes.

#### 2.4.1 Data cleaning and pre-processing

Of 2,768 participants who consented to anonymised data, 1,643 passed attention checks; a further 14 were excluded for falling outside the 18–40 age range, and 12 were excluded for having duplicated SONA IDs, leaving a final sample of 1,617. Given this age criterion, responses to education years and number of children (under and over 18) were screened for logical consistency with participant age (e.g., education years exceeding age minus four, or number of children over 18 exceeding what is biologically plausible given the minimum reported age of first birth). Inconsistent values were recoded as missing (education years, *n* = 138; children under 18, *n* = 37; children over 18, *n* = 36); extreme but plausible values were retained unaltered.

#### 2.4.2 Model fitting and comparison

For each outcome, we compared a reduced model with fixed effects of age, sex, and years of education (plus CM exposure for RQ2 and RQ3) against a full model that included a random intercept for country/region, testing whether between-country/region variability improved model fit. Model comparisons used likelihood ratio tests and change in log-likelihood. ICC quantified the proportion of variance attributable to country/region, and marginal and conditional R^2^ captured variance explained by fixed effects alone versus fixed and random effects combined. This framework was applied separately to CM scores (by type) and to perceived acceptability of CM (also by type, controlling for age, sex, and CM exposure), with the random intercept testing whether countries/regions differed at baseline after accounting for individual-level factors.

To test whether CM-outcome associations varied by country/region, separate models were estimated for psychosocial adjustment outcomes (post-traumatic growth, resilience, social support) and negative mental health outcomes (traumatic stress symptoms, psychological distress). Random slopes for CM exposure by country/region did not converge, so we adopted a fixed-effects interaction approach: for each outcome, a reduced model (age, sex, education, country/region, and CM subtypes as fixed effects) was compared against a full model adding CM subtype by country/region interaction terms. Type III tests checked each interaction for isolated effects despite a non-significant overall block.

## 3. Results

Participants had a median age of 22 years (range 18—40), a median of 14 years of formal education (range 0—30), and between 0 and 21 children under 18 and between 0 and 5 children over 18. Most (78.97%) identified as women; 18.86% as men, and 1.98% as non-binary (3 participants [0.18%] declined to answer). Most were single (54.17%), followed by long-term relationship (20.10%) or married (19.05%). Most were students or in full-time education (52.50%), 22.45% were employed full-time (≥ 35 hours/week), 12.12% were employed part-time (< 35 hours/week), and 5.57% unemployed. Demographics and raw mean scores by country appear in Tables S3 and S4.

### 3.1 Child maltreatment by country and region

Country/region-level clustering influenced most, but not all, forms of maltreatment. Likelihood ratio tests showed improved fit when country/region effects were included for physical neglect (ΔlogLik = 6.12), physical abuse (ΔlogLik = 14.69), emotional maltreatment (ΔlogLik = 21.45), and sexual abuse (ΔlogLik = 8.69), but not for witnessing domestic violence (ΔlogLik = 0.01; Table 1; Figure 1). The magnitude of this clustering also varied: emotional maltreatment showed the largest country/region-level variance (ICC = .06), followed by physical abuse (ICC = .04), sexual abuse (ICC = .03), and physical neglect (ICC = .02), while witnessing domestic violence showed essentially none (ICC < .01). Full models explained the most variance for emotional maltreatment (R²c = .09) and sexual abuse (R²c = .07), and the least for witnessing domestic violence and physical abuse (R²c = .02 and .05, respectively).

**Figure 1:**
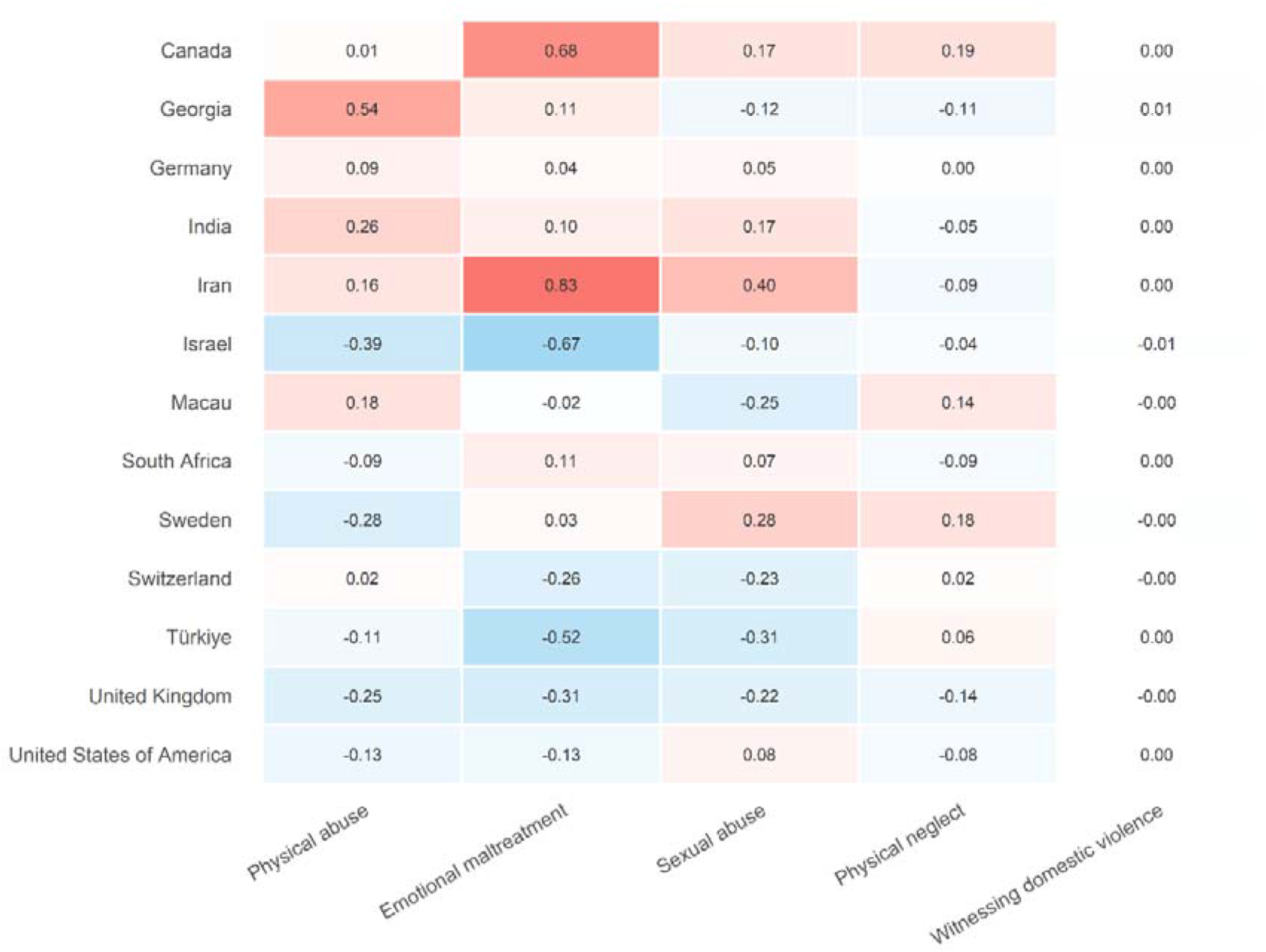
Country-adjusted mean CM subtype scores (physical abuse, emotional maltreatment, sexual abuse, physical neglect, witnessing domestic violence), controlling for age, gender, and education. Values are centred within subtype. Estimates reflect random-intercept country deviations from the grand mean for each CM subtype.

**Table 1:**
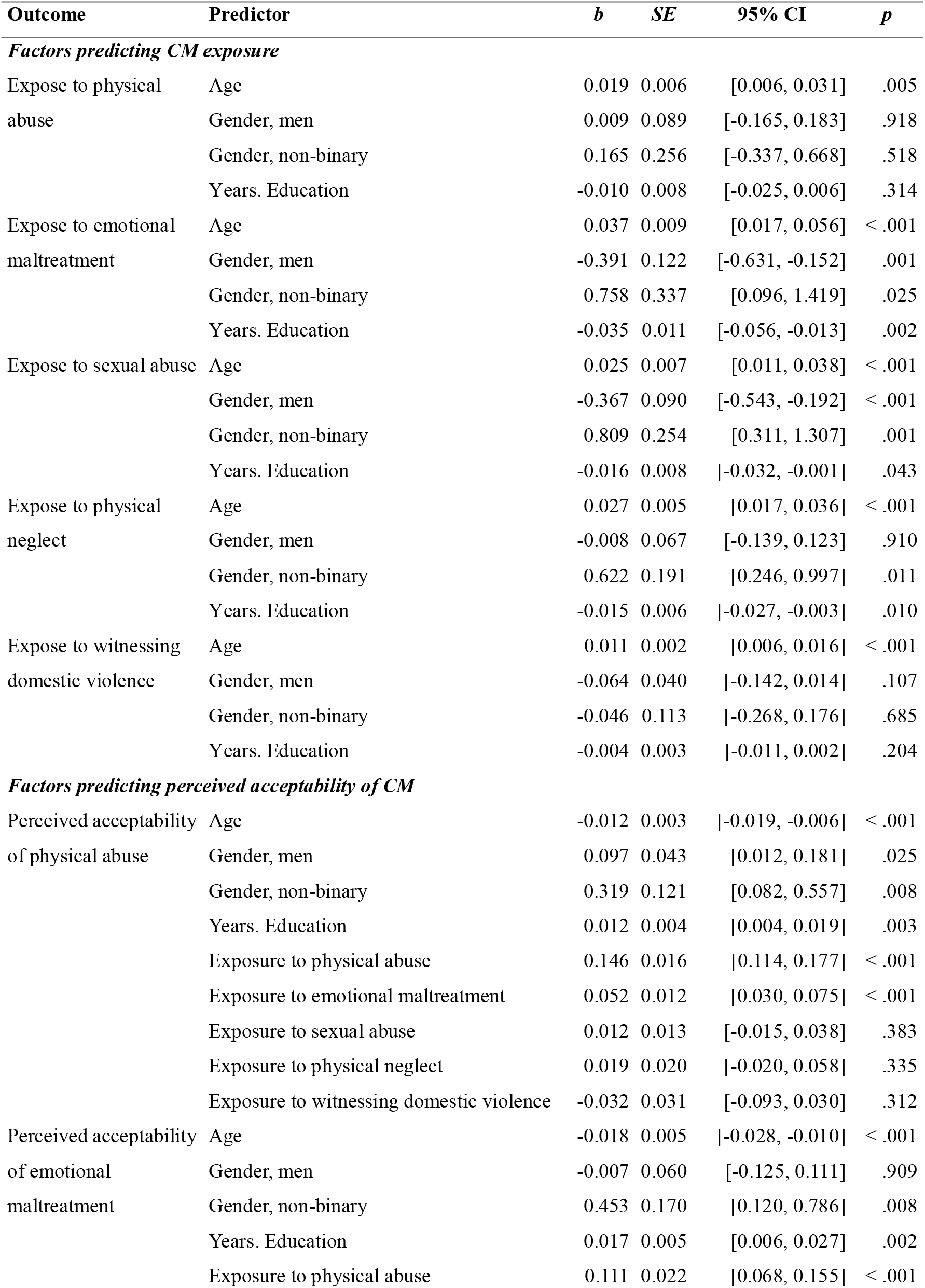
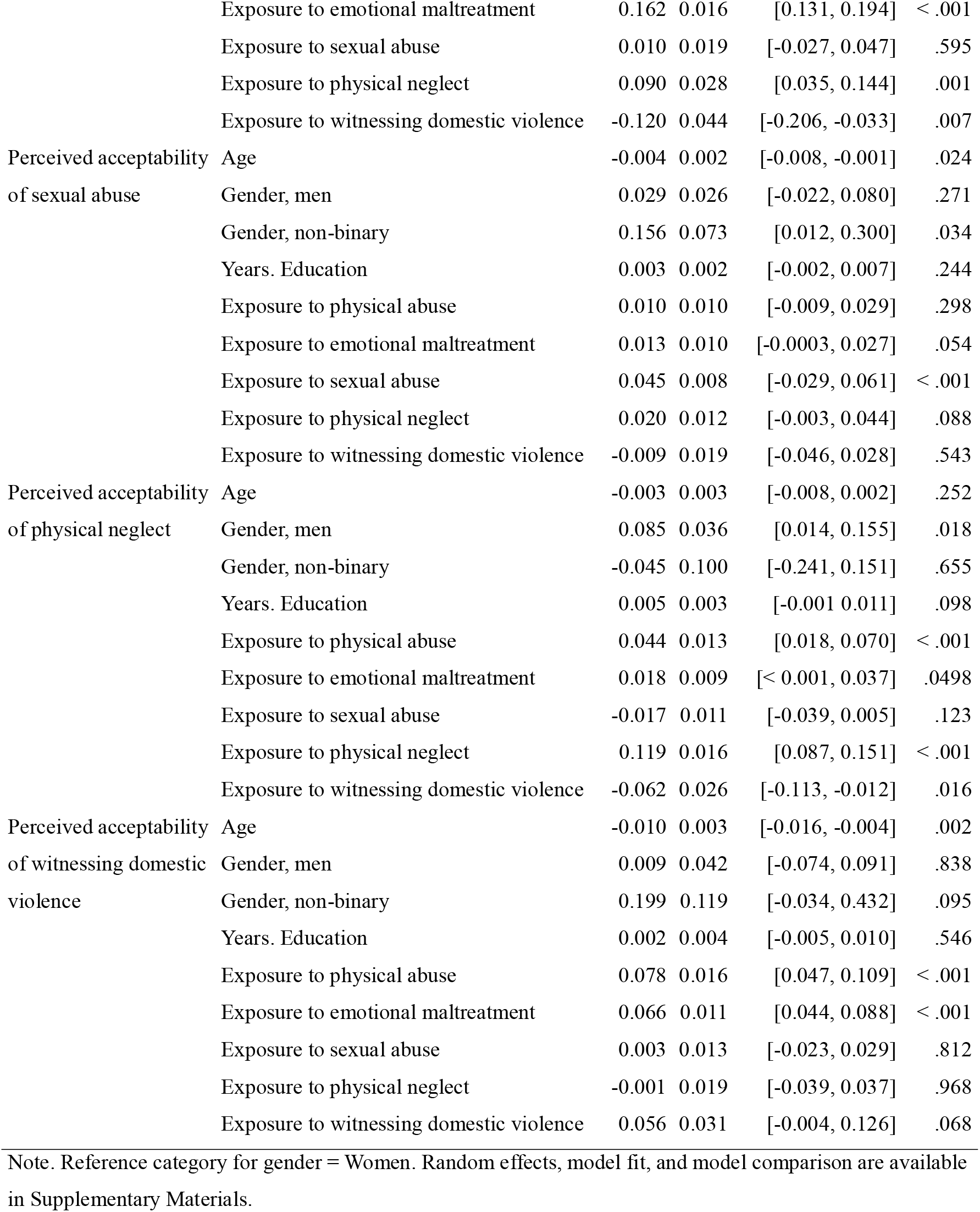
Factors predicting CM exposure and perceived acceptability.

**Table 2:**
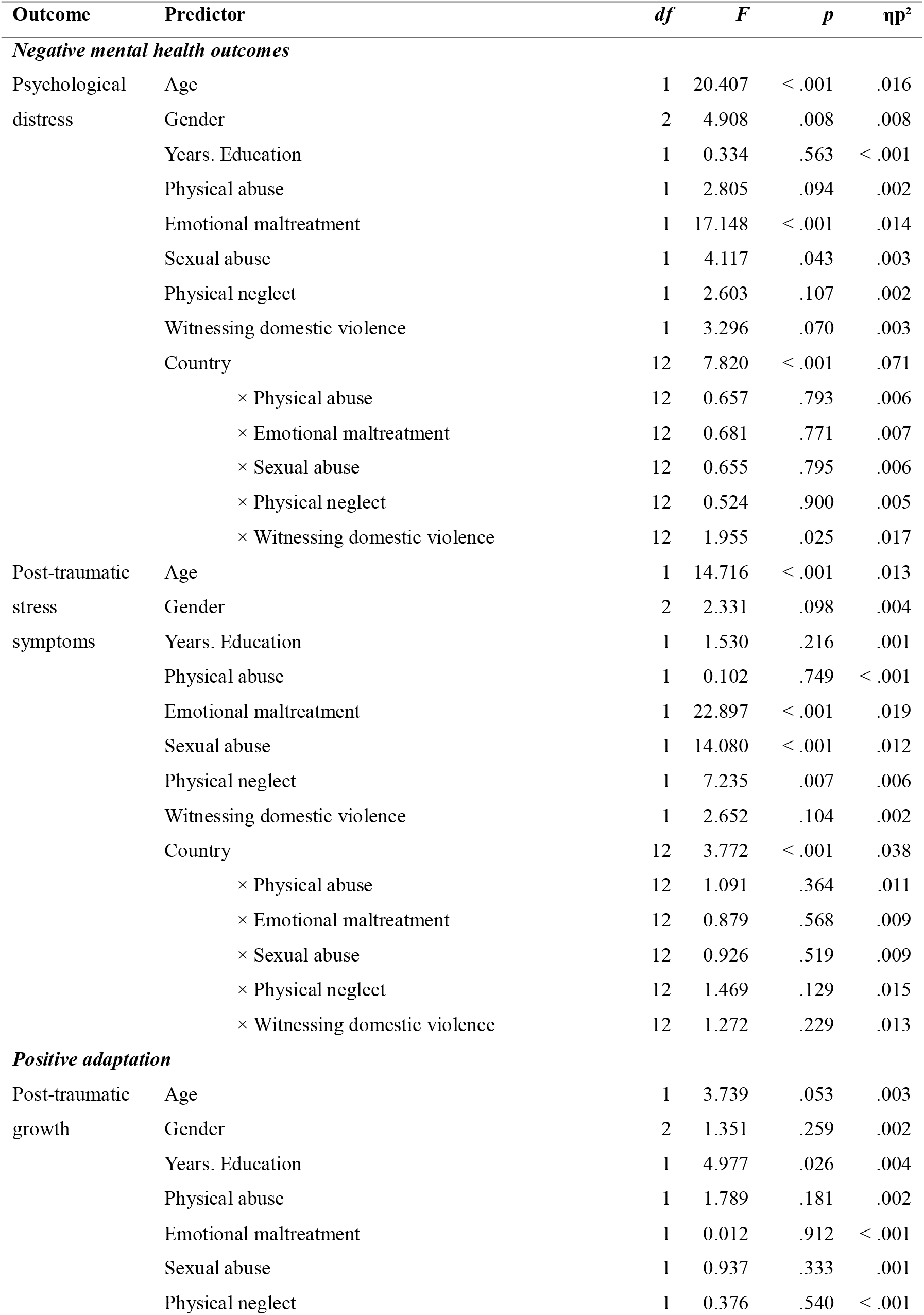
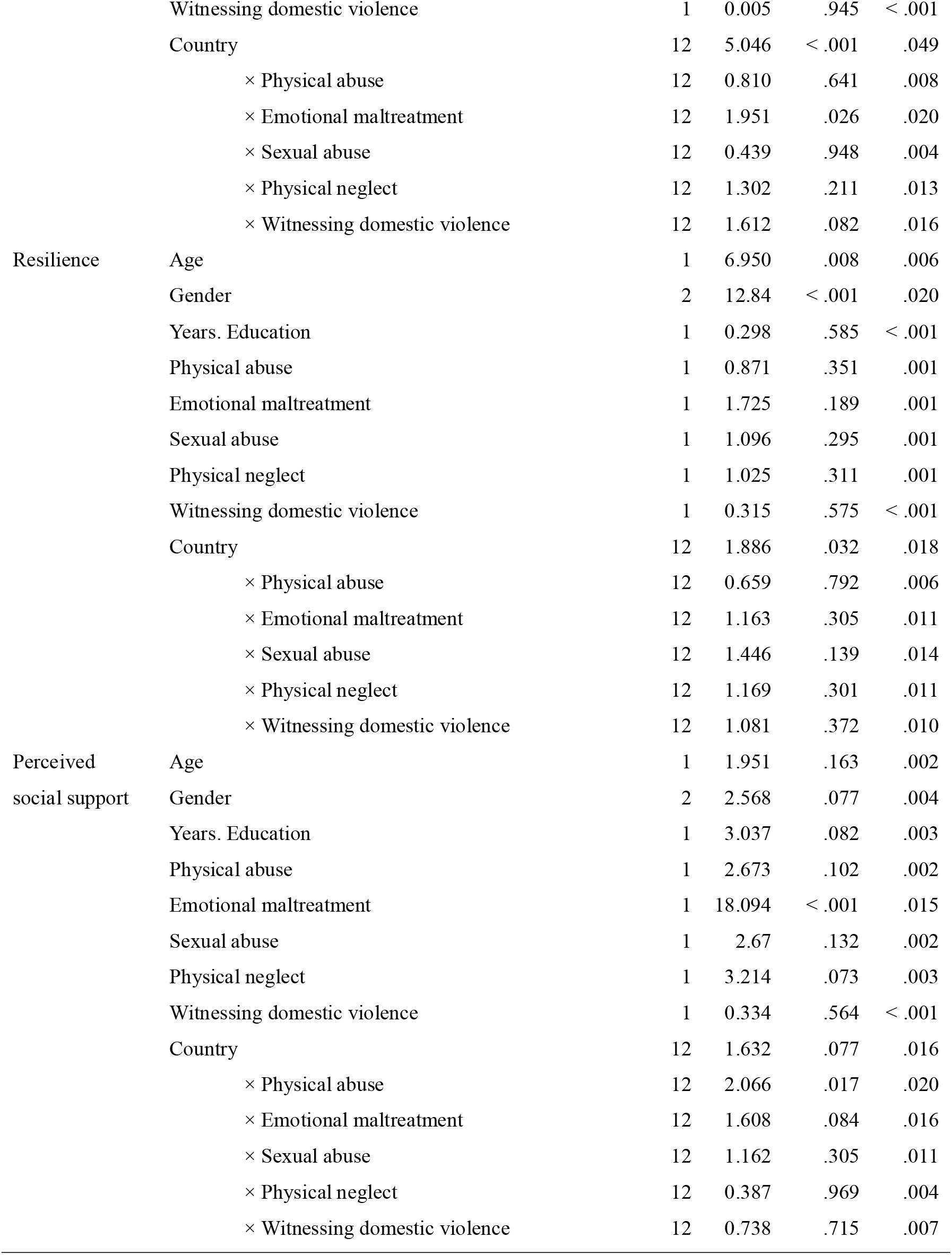
Associations between childhood maltreatment subtypes and negative mental health outcomes and positive adaptation across countries/regions.

Age showed the most consistent effect across all five outcomes: older participants reported more exposure to physical neglect (*b* = 0.03, *SE* = .01, *p* < .001), witnessing domestic violence (*b* = 0.01, *SE* = .002, *p* < .001), physical abuse (*b* = 0.02, *SE* = 0.01, *p* = .004), emotional maltreatment (*b* = 0.04, *SE* = .01, *p* < .001), and sexual abuse (*b* = 0.02, *SE* = .01, *p* < .001).

Gender effects, by contrast, were selective. Non-binary participants reported greater exposure than women for physical neglect (*b* = 0.62, *SE* = 0.19, *p* = .001), emotional maltreatment (*b* = 0.76, *SE* = 0.34, *p* = .025), and sexual abuse (*b* = 0.81, *SE* = .25, *p* = .001), but not for physical abuse or witnessing domestic violence (all *p* ≥ .518). Men, meanwhile, reported lower exposure than women to emotional maltreatment (*b* = −0.39, *SE* = .12, *p* = .001) and sexual abuse (*b* = −0.37, *SE* = .09, *p* < .001), with no significant difference for the other three outcomes.

Education followed a similar selective pattern to the non-binary gender effect: more years of formal education were associated with lower reported exposure to physical neglect (*b* = −0.02, *SE* = 0.01, *p* = .010), emotional maltreatment (*b* = −0.03, *SE* = .01, *p* = .002), and sexual abuse (*b* = −0.02, *SE* = .01, *p* = .043), but showed no association with physical abuse or witnessing domestic violence (all *p* ≥ .204).

### 3.2 Perceived acceptability of child maltreatment

Country/region effects improved model fit for all five forms of perceived acceptability (physical neglect ΔlogLik = 6.25; witnessing domestic violence ΔlogLik = 28.24; physical abuse ΔlogLik = 3.80; emotional maltreatment ΔlogLik = 28.21; sexual abuse ΔlogLik = 6.88), confirmed by likelihood ratio tests (Table 1; Figure 2). The degree of country/region-level clustering varied by outcome: physical abuse (ICC = .09) and emotional maltreatment witnessing and domestic violence (both ICC = .07) had the highest, while physical neglect and sexual abuse showed the least (both ICC = .03). Full models explained the most variance for emotional maltreatment (R²c = .26) and physical abuse (R²c = .23), and the least for physical neglect (R²c = .11) and sexual abuse (R²c = .08); witnessing domestic violence fell in between (R²c = .18).

**Figure 2:**
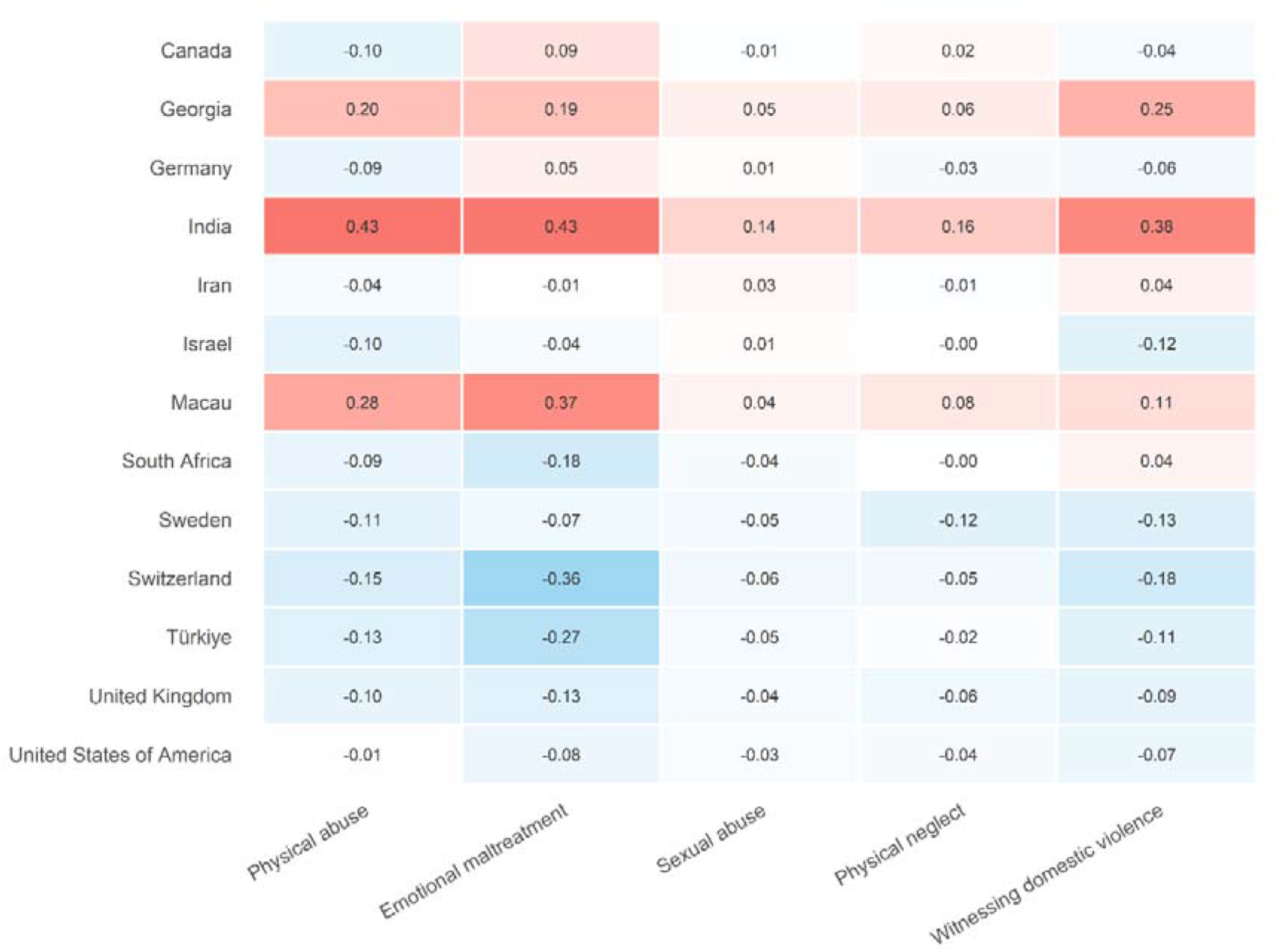
Country-adjusted mean perceived acceptability scores for each CM subtype (physical abuse, emotional maltreatment, sexual abuse, physical neglect, witnessing domestic violence), controlling for age, gender, education, and respondents’ own CM exposure across all five subtypes. Values are centred within subtype. Estimates reflect random-intercept country deviations from the grand mean for each acceptability measure.

**Figure 3:**
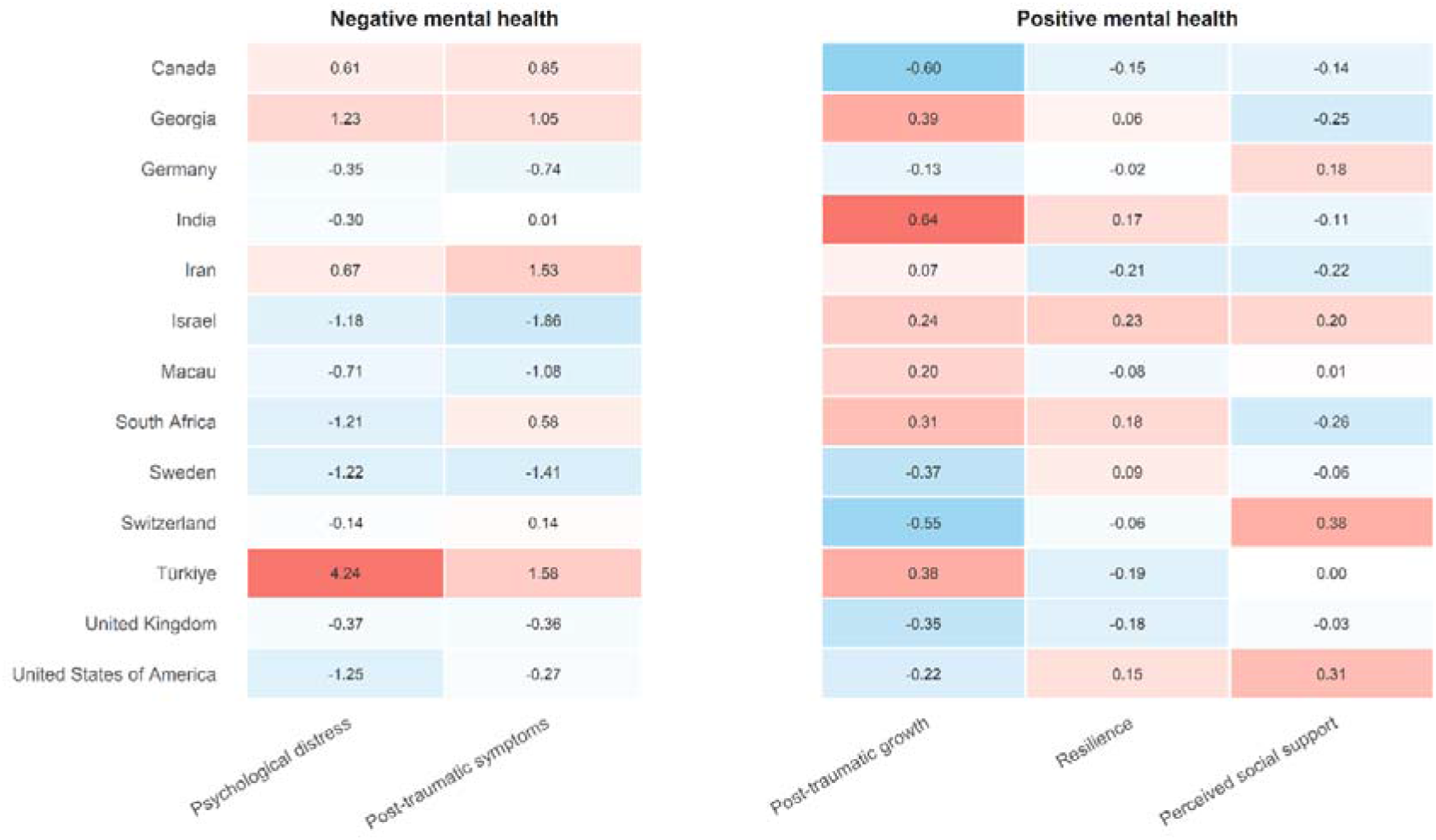
Country-adjusted mean scores for negative mental health outcomes (psychological distress and post-traumatic symptoms; right) and positive mental health outcomes (post-traumatic growth, resilience, and perceived social support; left), controlling for age, gender, education, and CM subtype scores. Values are centred within outcome, and colour scales are set separately for each panel. These estimates reflect country differences in outcome level; tests of whether the association between CM and each outcome varied by country are reported in Table 2.

A consistent pattern emerged across models: participants who had personally experienced more of a given maltreatment type tended to perceive that same type as more acceptable: physical neglect (*b* = 0.12, *SE* = 0.02, *p* < .001), physical abuse (*b* = 0.15, *SE* = 0.02, *p* < .001), emotional maltreatment (*b* = 0.16, *SE* = 0.02, *p* < .001), and sexual abuse (*b* = 0.04, *SE* = 0.01, *p* < .001). This pattern also extended across types: physical abuse exposure was associated with higher perceived acceptability of neglect (*b* = 0.04, *SE* = 0.01, *p* < .001), witnessing domestic violence (*b* = 0.08, *SE* = 0.02, *p* < .001), and emotional maltreatment (*b* = 0.11, *SE* = 0.02, p < .001); emotional maltreatment exposure was similarly associated with higher acceptability of neglect (*b* = 0.02, *SE* = 0.01, *p* = .0498), witnessing domestic violence (*b* = 0.07, *SE* = 0.01, *p* < .001), and physical abuse (*b* = 0.05, *SE* = 0.01, *p* < .001); and physical neglect exposure was associated with higher acceptability of emotional maltreatment (*b* = 0.09, *SE* = 0.03, *p* = .002). Witnessing domestic violence was the exception to this pattern: greater exposure was associated with lower perceived acceptability of both physical neglect (*b* = −0.06, *SE* = 0.03, *p* = .016) and emotional maltreatment (*b* = −0.12, *SE* = 0.04, *p* = .007).

Age effects pointed in a consistent direction across outcomes where a significant relationship emerged: older participants perceived witnessing domestic violence (*b* = −0.01, *SE* = .003, *p* = .002), physical abuse (*b* = −0.01, *SE* = 0.003, *p* < .001), emotional maltreatment (*b* = −0.02, *SE* = .01, *p* < .001), and sexual abuse (*b* = −.004, *SE* = .002, *p* = .023) as less acceptable than younger participants, though the effect for sexual abuse was negligible in size. Age showed no association with acceptability of physical neglect (*p* = .252).

Gender effects were selective. Men perceived greater acceptability than women for physical neglect (*b* = 0.08, *SE* = 0.04, *p* = .018) and physical abuse (*b* = 0.10, *SE* = 0.04, *p* = .025), with no significant effect for the remaining outcomes (all *p* ≥ .271). Non-binary participants perceived greater acceptability than women for physical abuse (*b* = 0.32, *SE* = 0.12, *p* = .008), emotional maltreatment (*b* = 0.45, *SE* = 0.17, *p* = .008), and sexual abuse (*b* = 0.16, *SE* = 0.07, *p* = .033), but not for physical neglect or witnessing domestic violence (both *p* ≥ .068).

Education showed a narrower effect: more years of formal education were associated with greater perceived acceptability of physical abuse (*b* = 0.01, *SE* = .004, *p* = .003) and emotional maltreatment (*b* = 0.02, *SE* = 0.01, *p* = .002), with no association for the other three outcomes (all *p* ≥ .098).

### 3.3 Child maltreatment and negative mental health outcomes

To test whether the association between childhood maltreatment and psychological distress varied by country/region, CM subtype by country interaction terms were added to a model containing country as a main effect. Overall, this interaction block did not significantly improve model fit, *F*(60, 1237) = 1.01, *p* = .459, indicating no strong evidence that the association between CM and psychological distress varied systematically by country/region. Examining the interaction terms by CM subtype, only the witnessing domestic violence by country interaction reached significance, *F*(12, 1237) = 1.96, *p* = .025; interactions involving physical neglect, physical abuse, emotional maltreatment, and sexual abuse did not (all *p* ≥ .771).

The same approach was applied to post-traumatic stress symptoms. The CM subtype by country interaction block did not significantly improve model fit, *F*(60, 1161) = 1.27, *p* = .086, and no individual CM subtype by country interaction reached significance (all *p* ≥ .129).

### 3.4 Child maltreatment and psychosocial adjustment

The same interaction approach was applied to resilience. The CM subtype-by-country interaction block did not significantly improve model fit, *F*(60, 1229) = 1.23, *p* = .118, and no individual CM subtype-by-country interaction reached significance (all *p* ≥ .139).

For post-traumatic growth, the CM subtype by country interaction block also did not significantly improve model fit, *F*(60, 1168) = 1.21, *p* = .135. Examining the interaction terms by CM subtype, only the emotional maltreatment by country interaction reached significance, *F*(12, 1168) = 1.95, *p* = .025; interactions involving physical neglect, physical abuse, sexual abuse, and witnessing domestic violence did not (all *p* ≥ .082).

For perceived social support, the CM subtype-by-country interaction block did not significantly improve model fit, *F*(60, 1216) = 1.17, *p* = .184. Examining the interaction terms by CM subtype, only the physical abuse (*F*(12, 1216) = 2.07, *p* = .017) by country interaction reached significance; interactions involving physical neglect, emotional maltreatment, sexual abuse, and witnessing domestic violence did not (all *p* ≥ .083).

Across all five outcomes, the overall test of country-level moderation was non-significant, indicating that CM-psychosocial associations were largely consistent across countries/regions rather than culturally specific. Isolated CM subtype by country interactions emerged for three of the five outcomes (witnessing domestic violence for psychological distress; emotional maltreatment for post-traumatic growth; physical abuse for perceived social support) although none were accompanied by a significant omnibus test.

## 4. Discussion

This international cross-sectional study was conducted across 13 countries and regions, encompassing diverse cultural, linguistic, and socio-economic contexts. Differences emerged between countries and regions in CM scores and perceived acceptability, with both patterns varying across CM types. By contrast, associations between CM and negative mental health outcomes and positive adaptation were largely consistent across countries and regions with only isolated, subtype-specific exceptions. Our samples were not representative of the general populations in each country and region so cross-country differences should be interpreted as differences across recruited samples rather than population-level estimates. Nevertheless, the consistency of overall patterns supports the broader relevance of our findings while underscoring the need for future population-based studies to confirm their generalisability.

Across countries and regions, self-reported CM showed significant between-country/region variability across most types, particularly for emotional maltreatment (highest among Iranian participants, lowest among Israeli participants). Witnessing domestic violence showed negligible differences. These variations align with prior findings (Wadji *et al*. 2023b), and mechanisms underlying them (e.g., social norms, parenting practices, service accessibility) should be a focus of future research.

### 4.1 National and regional differences in perceived acceptability of child maltreatment

Perceived acceptability of CM also varied significantly across countries and regions, consistent with our pilot study (Wadji *et al*. 2023b), with the largest effect observed for emotional maltreatment. A similarly pronounced difference emerged between Switzerland and India, a WEIRD and a non-WEIRD country with markedly different legal and sociocultural frameworks (Fernandes *et al*. 2021; Jud and Knüsel 2019), consistent with the idea that culture shapes parenting practices and associated social norms (Bornstein 2013; Lansford 2010), as reflected in child protection laws (Zolotor and Puzia 2010). However, WEIRD countries did not consistently rank higher or lower in CM acceptability than non-WEIRD countries, contradicting Hayes and O’Neal’s (2018) finding that members of more survival-oriented cultures show lower tolerance for CM. A critical next step is to investigate the factors underlying these cross-cultural differences and whether culture-specific profiles distinguish physical from psychological violence, or threat-based (abuse) from deprivation-based (neglect) experiences (Lee *et al*. 2014; McLaughlin *et al*. 2014).

Greater exposure to CM was associated with increased perceived acceptability of CM with broader cross-type associations for physical neglect, physical abuse, and emotional maltreatment, and a type-specific association for sexual abuse. This extends prior findings confined largely to neglect and sexual abuse (Bartoli *et al*. 2024; Wadji *et al*. 2023b), suggesting that CM experiences may shape wider normative beliefs about caregiving through normalisation, desensitisation, or adaptation to adverse environments (Corso *et al*. 2025; Kawaguchi *et al*. 2025); a concern given the potential for intergenerational continuity of CM (Langevin *et al*. 2026).

Men generally perceived physical abuse as more acceptable than women, consistent with prior evidence of greater acceptance of corporal punishment among men (Lansford *et al*. 2020). Education showed a more nuanced relationship: in line with prior work linking parental education to reduced maltreatment risk (Lansford *et al*. 2015), higher educational attainment in our sample was associated with lower reported exposure to physical neglect, emotional maltreatment, and sexual abuse, but with greater perceived acceptability of physical abuse and emotional maltreatment, contradicting literature linking education to lower endorsement of corporal punishment (Hendaus *et al*. 2020; Klevens *et al*. 2019; Taylor *et al*. 2011) though echoing a smaller body of work reporting null or contradictory education-attitude associations (Landa-Blanco *et al*. 2026; Yıldız and Ceyhan 2025). This suggests the education-acceptability link found elsewhere might not generalise across our culturally diverse sample where attitudes may be shaped more by personal experiences or prevailing social norms than education alone (Landa-Blanco *et al*. 2026). Further research should test whether this reflects sample characteristics or a replicable finding.

### 4.2 Child maltreatment, negative mental health outcomes, and positive adaptation

Greater exposure to CM was generally associated with poorer mental health, although not uniformly across subtypes. Emotional maltreatment and sexual abuse were independently associated with higher psychological distress, and emotional maltreatment, sexual abuse, and physical neglect with more severe PTSD symptoms. Physical abuse and witnessing domestic violence showed no significant independent association with either outcome. This divergence warrants further study, particularly for witnessing domestic violence given its well-documented negative impact on mental health (Jagasia *et al*. 2024; Mueller and Tronick 2019). Cross-national variation in these associations was limited: the overall CM subtype by country/region interaction did not significantly improve model fit for either outcome, indicating consistent association strength across countries/regions with only an isolated exception for witnessing domestic violence and psychological distress. Our findings support the notion that CM’s negative repercussions on mental health are a global phenomenon (Carr *et al*. 2020a), while cultural factors shape the psychological processing and expression of traumatic distress (Schnyder *et al*. 2016), including cross-cultural differences in self-stigmatisation, meaning-making, and emotional processing, all of which may explain variability in symptom expression (Hashim *et al*. 2026).

Associations with CM exposure were more limited for positive adaptation. No CM subtype showed a significant independent association with resilience, only emotional maltreatment was independently associated with lower perceived social support, and post-traumatic growth showed no significant association with any subtype. As with negative outcomes, cross-national variation was limited: the overall interaction block was non-significant for all three outcomes, with only isolated exceptions for emotional maltreatment (post-traumatic growth) and both physical abuse and emotional maltreatment (perceived social support). Where associations did emerge, they suggested CM may compromise core psychological and interpersonal resources supporting adaptive functioning (e.g., emotion regulation, self-efficacy, trust in others) that develop within caregiving environments often disrupted by abuse and neglect (Fares-Otero *et al*. 2025). That emotional maltreatment was not associated with greater post-traumatic growth in this larger, more culturally diverse sample contrasts with pilot findings (Wadji *et al*. 2023a), suggesting positive adaptation may be less consistently observed than maladaptive outcomes and more dependent on contextual and cultural factors shaping meaning-making and recovery (Yoon *et al*. 2021, 2023).

### 4.3 Strengths and limitations

Strengths of this study include the large, multinational sample spanning 13 countries and regions, harmonised assessment procedures across sites, and simultaneous examination of CM exposure, perceived acceptability, and psychosocial outcomes. Limitations include the cross-sectional design which precludes accounting for intervening events (e.g., additional trauma between childhood and assessment) or current contextual factors such as living in conflict-affected regions. Retrospective self-report measures are also susceptible to recall bias (Schmidt *et al*. 2023) and many measures lack published translations in target countries. Future research should assess participants routinely to mitigate recall bias and work towards standardised, cross-culturally validated CM measures (Fares-Otero and Seedat 2024). Finally, recruitment strategies varied across sites (social media, personal/professional networks, university subject pools), introducing heterogeneity which future work should aim to address using representative samples within each country/region.

### 4.4 Conclusion

This cross-cultural study of adults across 13 countries and regions demonstrates that CM types vary in prevalence, perceived acceptability, and associations with negative and positive mental health outcomes. Exposure to specific CM types also appears to be linked to perceived acceptability of other subtypes, highlighting both type-specific and general dimensions of family violence. These findings underscore the need for assessment and interventions that account for cultural and type-specific differences, with future resource developing validated, culture-specific measures and examining moderators and mediators of acceptability and adaptation using longitudinal methods.

## Author contributions

- D Jenkins: conceptualisation, data curation, formal analysis, investigation (South Africa, Switzerland), methodology, project administration, resources, visualisation, writing (original draft), writing (review and editing)
- R Langevin: investigation (Canada), resources, writing (original draft), writing (review and editing)
- S Rita Balle: investigation (Germany), resources, writing (original draft), writing (review and editing)
- O Jiaqing: investigation (Macau), resources, writing (original draft), writing (review and editing)
- S Porat-Butman: investigation (Israel), resources, writing (original draft), writing (review and editing)
- E Levi-Gigi: investigation (Israel), resources, writing (review and editing)
- G Ayas: investigation (Türkiye), resources, writing (review and editing)
- D Ceylan: investigation (Türkiye), writing (review and editing)
- N Fares-Otero: writing (original draft), writing (review and editing)
- M Fernández Boente: investigation (Switzerland), writing (review and editing)
- S Halligan: investigation (UK), resources, writing (review and editing)
- B Jansson: investigation (Sweden), writing (review and editing)
- S Kamari Songhorabadi: investigation (Iran), resources, visualisation, writing (review and editing)
- G Khayati: investigation (Iran)
- F Khormaei: investigation (Iran)
- C Martin-Sölch: investigation (Switzerland), resources, writing (review and editing)
- M Pfaltz: investigation (Sweden), resources, writing (review and editing)
- M Piccolo: investigation (USA), resources, writing (review and editing)
- V Şar: investigation (Türkiye), resources, writing (review and editing)
- U Schnyder: supervision, writing (review and editing)
- S Seedat: investigation (South Africa), resources, writing (review and editing)
- T Tandon: investigation (Switzerland), writing (review and editing)
- T Türk Kurtça: investigation (Türkiye), writing (review and editing)
- E Bartoli: conceptualisation, methodology, project administration, supervision, writing (original draft), writing (review and editing)
- G Spies: conceptualisation, investigation (South Africa), methodology, project administration, resources, supervision, writing (original draft), writing (review and editing)
- N Iqbal: conceptualisation, investigation (India), methodology, project administration, resources, supervision, writing (original draft), writing (review and editing)
- M Oe: conceptualisation, methodology, project administration, resources, supervision, writing (original draft), writing (review and editing)

## Ethics statements

Umbrella ethics clearance for this study was provided by the Stellenbosch University Health Research Ethics Committee (ref: N23/03/026). Additionally, the following local ethics committees provided ethics clearance for data collection in their countries/regions: McGill University Ethics Board Office for Canada (ref: 23-08-039), Ilia State University Faculty of Sciences and Arts Research Projects Ethics Review Commission for Georgia (ref: IL24/02/11), Jamia Millia Islamia Department of Psychology Ethnical Committee for India (date: 07-11-2023), Research Ethics Committee of Shiraz University for Iran (ref: IR.US.REC.1401.011), Bar-Ilan University IRB Committee for Israel (ref: 2505261121), Research Ethics Panel at the Department of Psychology, Faculty of Social Sciences, University of Macau for Macau (ref: 2024-08), Mid Sweden University Ethics Review Authority for Sweden (ref: 2023-06703-01), University of Fribourg Internal Review Board of the Department of Psychology for Switzerland (ref: 2024-968), Koç University Social Sciences Research Ethics Committee for Türkiye (ref: 2023.297.IRB3.140), and Harvard University Institutional Review Board for USA (ref: IRB24-0420). Ethics exemption for data collection in Germany was provided by Bundeswehr Universität München (date: 12-07-2023).

## Data availability

Materials that support the findings of this study are openly available in Zenodo at http://doi.org/10.5281/zenodo.23191665. Free-form survey fields (e.g., ‘other’ columns on the demographics survey) have been removed to protect participant confidentiality but are available from the corresponding author on reasonable request.

