## Supplementary Materials for "When Is Maltreatment ‘Acceptable’? Cross-Cultural Patterns in Child Maltreatment Exposure and its Perceived Social Acceptability Across 13 Countries and Regions"

### Random effects, model fit, and model comparison for Table 1

CM Exposure:

- Physical abuse: σ^2^_country_ = 0.07, σ^2^_residual_ = 1.73, ICC = .04, R^2^_m_ = .01, R^2^_c_ = .05, χ^2^(1) = 29.39, p < .001, ΔlogLik = 14.69
- Emotional maltreatment: σ^2^_country_ = 0.19, σ^2^_residual_ = 3.19, ICC = .06, R²_m_ = .03, R²_c_ = .09, χ^2^(1) = 42.90, p < .001, ΔlogLik = 21.45
- Sexual abuse: σ^2^_country_ = 0.06, σ^2^_residual_ = 1.76, ICC = .03, R^2^_m_ = .03, R^2^_c_ = .07, χ^2^(1) = 17.39, p < .001, ΔlogLik = 8.69
- Physical neglect: σ^2^_country_ = 0.02, σ^2^_residual_ = 0.97, ICC = .02, R^2^_m_ = .04, R^2^_c_ = .06, χ^2^(1) = 12.25, p < .001, ΔlogLik = 6.12
- Witnessing domestic violence: σ^2^_country_ = 0.0002, σ^2^_residual_ = 0.36, ICC < .001, R^2^_m_ = .02, R^2^_c_ = .02, χ^2^(1) = 0.02, p = .894, ΔlogLik = 0.01

Perceived acceptability of CM:

- Physical abuse: σ^2^_country_ = 0.03, σ^2^_residual_ = 0.36, ICC = .09, R^2^_m_ = .16, R^2^_c_ = .23, χ^2^(1) = 65.60, p < .001, ΔlogLik = 32.80
- Emotional maltreatment: σ^2^_country_ = 0.06, σ^2^_residual_ = 0.71, ICC = .07, R^2^_m_ = .20, R^2^_c_ = .26, χ^2^(1) = 56.43, p < .001, ΔlogLik = 28.21
- Sexual abuse: σ^2^_country_ = 0.004, σ^2^_residual_ = 0.13, ICC = .03, R^2^_m_ = .06, R^2^_c_ = .08, χ^2^(1) = 13.76, p < .001, ΔlogLik = 6.88
- Physical neglect: σ^2^_country_ = 0.01, σ^2^_residual_ = 0.25, ICC = .03, R^2^_m_ = .09, R^2^_c_ = .11, χ^2^(1) = 12.50, p < .001, ΔlogLik = 6.25
- Witnessing domestic violence: σ^2^_country_ = 0.03, σ^2^_residual_ = 0.35, ICC = .07, R^2^_m_ = .11, R^2^_c_ = .18, χ^2^(1) = 56.48, p < .001, ΔlogLik = 28.24
